# Mental Health Problems in Nepalese Migrant Workers and their Families

**DOI:** 10.1101/2020.08.04.20168104

**Authors:** Pashupati Mahat, Kevan Thorley, Karuna Kunwar, Smriti Ghirime

## Abstract

**Background:** Nepal has an economy increasingly dependent on remittences from migrant wokers. Mental health problems affect a significant number of these workers and the prevalence of mental health problems in the left behind families of migrant workers is high. Facilities for the psychosocial support of migrant workers and their families are scarce. A project to provide such support is described.

**Objective:** We aim to describe the mental health problems of Nepalese migrant workers and their family members remaining at home in Nepal.

**Methods:** Families of migrant workers from nine project districts were interviewed and offered appropriate psychosocial counseling. The psychosocial problems experienced by families left behind in Nepal (women, children and elderly parents) were assessed.

**Results:** Social isolation, excessive worry, low mood, fearfulness and sleep disturbances were frequently reported. Wives and mothers of migrant workers experienced anxiety, depression and suicidal ideation as well as suicide attempts.. Domestic violence, death of migrant workers, health problems of migrant workers and their families and the difficulties of communication when working overseas were found to be contributing factors for psychosocial and mental health problems.

## Background

Labour migration has become common practice among Nepalese youth and the established main source of earning in many families and has contributed to 29.4 percent of GDP in 2015 indicating a national shift towards a remittance dependent economy from traditional agriculture. (Baruah 2018)

Migration has significant social costs. The separation of families causes social and psychological problems among family members left behind, especially women and children as they face the problems of social isolation, abuse and harassment, breakdown of relationships among family members and community and social stigma. (Helvetas Safer Migration Project 2019) Children of migrant workers experience separation anxiety, low self-esteem, vulnerability to trafficking and abuse and social stigma.(Centre for Mental Health and Counselling Nepal 2015)

The Safer Migration project (SaMi) is a bilateral initiative of the Government of Nepal (GoN) and the Government of Switzerland. The project is implemented through a partnership between HELVETAS Swiss Interco-operation Nepal and the Ministry of Labour, Employment and Social Security. (Federal Department of Foreign Affairs 2019)

The Centre for Mental Health and Counseling Nepal (CMC-Nepal), national partner of SaMi project has supported the implementation of a program of psychosocial support targeting the families of migrant workers and distressed returnee migrants in nine project districts ranging from the hill regions to the plains of the Terai since 2013.

## Objective

To describe the psychosocial issues experienced by returned migrant workers and the families of returned and absent migrant workers.

## Methods

The Psychosocial intervention component of the project included building capacity of staff of local partners of SaMi/HELVETAS. The first phase was a pilot implemented in two districts and expanded to seven others by June, 2016. CMC-N trained four staff in each district in psychosocial counselling during a six months practice based training program. 39 psychosocial counsellors were trained and working in nine districts by the end of June, 2016. Psychosocial counselling services were also provided through home visits.

Selection criteria for the sample population were returned migrant workers with psychosocial and mental health conditions and the families of migrant workers working in six GULF countries and Malaysia. We included wives, children and other family members (mothers, siblings) of migrant workers. The sample (beneficiaries of the project) were identified through the help of local returnee volunteers, school teachers, local media, the information and counselling center (ICC), women’s network groups (mother’s groups, women’s cooperatives) and the district police office.

The project attempted to address psychosocial problems through a service delivered by trained counselors who visited affected families, developed trust with individual clients and family members and explored the problems (mainly symptoms as complaints). The aim of the intervention was to reduce symptoms through strengthening the ability of clients to cope with their situations and feelings. The key psychosocial intervention skills adopted by counsellors were active listening skills, helping to analyse the consequence of the suffering of client, helping to change the focus of the client from symptoms and relationship problems to functional abilities and strengths, tips for self-care such as drinking water regularly, dietary advice, sleep hygiene and stress reducing breathing exercises and intervention focused questions.

Counsellors received a six month training course of 900 hours consisting of 440 hours theory and 460 hours supervised practice.. Counselling sessions were observed and coached Distance supervision was also provided. Trainee counsellors maintained case documentation including details of counselling session plans before the sessions with the help of an assigned psychologist supervisor.

Data for this study were collected by the counsellors working with returned workers and their families and entered into an excel spreadsheet.

## Findings

95 returned migrant workers and 652 family members of absent migrant workers received psychosocial services from 36 counsellors from July, 2015 to June 2016. 64/95 (67%) returnees were male and 31/95 (33%) female. 53/95 (56%) of these returnees suffered from anxiety, 22/95 suffered from (23%) depression, 6/95 (6%) had made suicide attempts or experienced suicidal ideation and 10/95 (11%) were found to have serious mental illness. (Table 1). The contributing factors reported were experiences work-related problems such as more arduous work than described in their initial contract, low pay, illegal status and violence (threat-verbal and in few cases physical assault). Experience of violence was reported by distressed female returnees in their work place, mainly by the domestic workers.

**Table 1.**
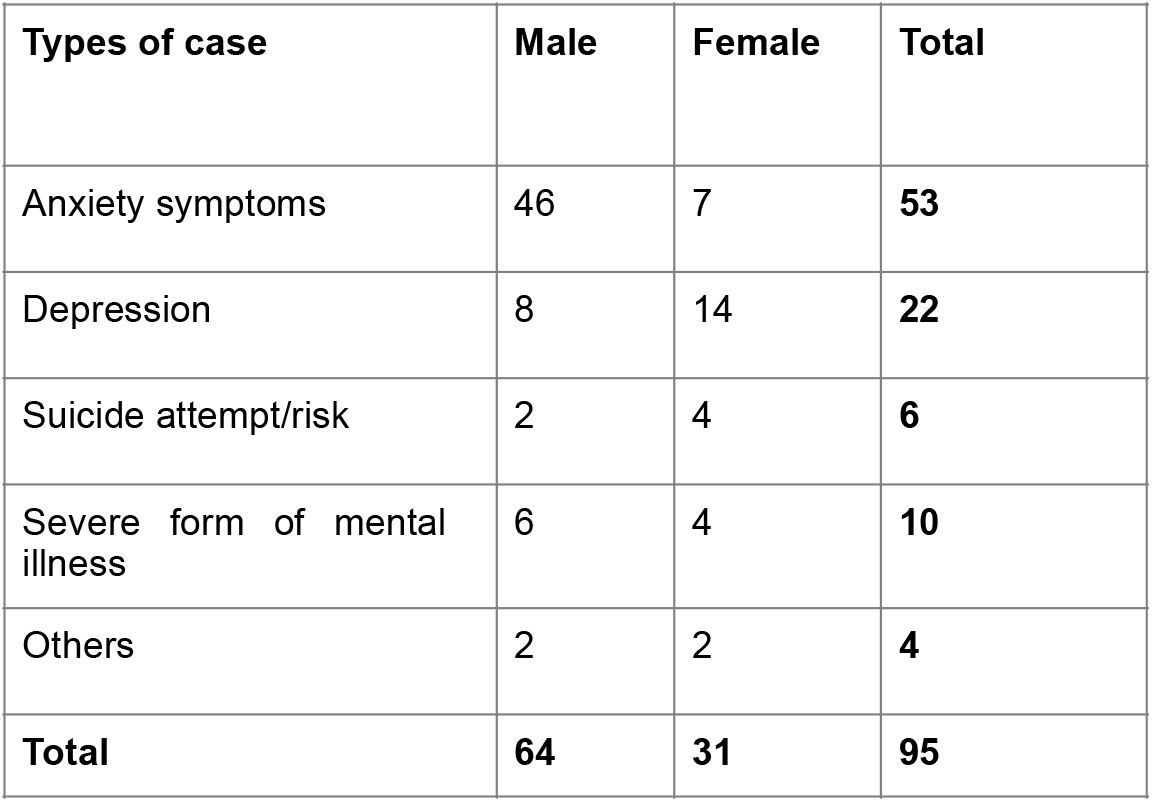
Psychosocial problems experienced by returnee migrant workers attending the psychosocial support program.

**Table 2.** Psychosocial problems experienced by left behind clients attending the psychosocial support program.

| **Types of case** | **Male** | **Female** | **Total** |
| --- | --- | --- | --- |
| Anxiety symptoms | 7 | 369 | **376** |
| Depression | 19 | 147 | **166** |
| Suicide attempt/risk | 3 | 41 | **44** |
| Severe form of mental illness | 2 | 9 | **11** |
| Others | 16 | 39 | **45** |
| **Total** | **47** | **605** | **642** |

The project supported 747 individual clients with a psychosocial counselling service in a one year period. 652/747 of these clients were left-behind family members. 605 /652 (93%) left-behind clients were female and 52/652 (8%) were male. 376/642 (56%) of left behind clients were suffering from anxiety. 166/642 (26%) suffered from depression., 44/642 (7%) expressed suicidal ideation or had attempted suicide, 11/642 (2%) had severe mental illness and 45/642 (7%) had other problems without medical diagnosis

The largest group of clients was composed of the wives of migrant workers (462/747 (62%)). Mothers of migrant workers made up 132/747/d (18%) and children 36/747(5%). (Table 3) The age distribution of clients is shown in table 4.

**Table 3.** Client’s relationship with migrant workers.

| **Relation** | **N** | **%** |
| --- | --- | --- |
| Wife | 462 | 62 |
| Mother | 132 | 18 |
| Father | 20 | 3 |
| Husband | 11 | 2 |
| Children | 36 | 5 |
| Other family member | 19 | 3 |
| Returnee MW | 67 | 9 |
| **Total** | **747** | **100** |

**Table 4.** Age distribution of clients attending psychosocial counselling.

| Age | Number of clients | % |
| --- | --- | --- |
| Under 16 | 44 | 6 |
| 17-25 | 157 | 21 |
| 26-35 | 286 | 38 |
| 36-45 | 124 | 17 |
| Over 45 | 136 | 18 |
| Total | 747 | 100 |

Domestic violence was the most frequently found contributory factor for psychosocial problems (anxiety, depression and suicide). Health problems of migrant workers, death of migrant worker and difficulties in communication with home for the migrant worker contributed to mental health problems (anxiety, depression and suicide) in the families of migrant workers. (Table 5)

## Discussion

The prevalence of both depression and anxiety in the families of migrant workers was found in this study to be twice that found in a study of the mental health of the general population of Nepal.(Rossi 2008) Returning migrant workers also reported higher levels of depression and anxiety than in the home population. We focused on migration to GULF countries and Malaysia the two areas to which Nepalese workers migrate most frequently.

A strength of the study is the large sample size, representing the families of migrant workers from a variety of geographical districts in Nepal. Psychosocial issues experienced by the families of migrant workers have not previously been studied in the context of Nepalese migration. The numbers of returning workers in the study, however are small, especially those of women. Problems of data collection in Nepal did not allow us to disaggregate some of the data relating to returning workers and left-behind clients so that analysis of contributory factors between these two groups is unsatisfactory. Our findings are consistent with those of studies of the mental health of left-behind families of migrant workers in other countries such as Sri Lanka and Jamaica. (Siriwardhana 2015, Jones 2008). Risal et al (2015) found that women in the general population of Nepal experience higher rates of depression than men. Studies in Sri Lanka where nearly ten percent of the adult population is employed abroad as migrant workers, demonstrate a prevalence of common mental disorder of 21% among spouses of migrant workers left behind and of 30% in non-spouse caregivers. (Siriwardhana 2015)

## Data Availability

The study was conducted in Nepal where facilities are basic. I have the relevant data in my personal files.

## Clinical Implications

Future research in this area may investigate further the work-related aspects of the mental health problems of returning migrant workers and possible preventive strategies for these workers and their left-behind family member.

## Ethical Approval

Ethical apporval for this study was granted by the Nepal Health Research Ethics Committee.

### Summary

What is already known about this subject?

- Nepal’s economy depends on migrant workers
- There is little provision for psychosocial support for these workers and their families
- The prevalence of mental health problems in returning workers is thought to be high

What are the new findings?

- Depression anxiety and relationship problems are frequent in returning workers and their families
- Provision of services may be made by co-operation between existing organisations
- The early ersults of such provision are positive

How might it impact on clinical practice in the future?

Provision of psychosocial support for such groups could ahve a beneficial effect on them and the economy.

## References

Baruah N, Arjal N, Nepalese Labor Migration—A Status Report. The Asia Foundation June 6, 2018. https://asiafoundation.org/2018/06/06/nepalese-labor-migration-a-status-report/ (accessed 16.4.2019)

Centre for Mental Health and Counselling Nepal Annual report, 2015, (pp-18-20)

Federal Department of Foreign Affairs FDFA, Swiss Agency for Development and Cooperation SDC Sami Safer Migration Project http://www.sami.org.np/ Accessed 9.4.2019

Helvetas Safer Migration Project https://www.helvetas.org/en/nepal/what-we-do/how-we-work/our-projects/Asia/Nepal/nepal-safer-migration (Accessed 16.4.2019)

Jones (2004), Children’s *Experiences of Separation from Parents as a Consequence of Migration.* Caribbean Journal of Social Work 3, p.89-109.

Risal A, Manandhar K, Linde M, Steiner TJ, Holen A. Anxiety and depression in Nepal: prevalence, comorbidity and associations BMC Psychiatry 2016 16:102 https://doi.org/10.1186/s12888-016-0810-0

Rossi, A (2008). The impact of migration on children in developing countries.” Papers of youth migration conferences (pp.24-26).

Siriwardhana C, Wickramage K, Siribaddana S, Vidanapathirana P, Jayasekara b, Weerawarna S, Pannala G, Adikari A Jayawaweera K, Pieris S, Smathipatla A Common mental disorders among adult memebrs of ‘left behind’ international migrant worker families in Sri Lanka BMC Public Health 2015 Mar 28; 15:200 doi 10.1186/s12889-015

